# Advancing In-Hospital Mortality Prediction for Acute Myocardial Infarction: an analysis from the American Heart Association Get-With-the-Guidelines Coronary Artery Disease Registry

**DOI:** 10.1101/2025.07.02.25330775

**Authors:** Juan Zhao, Haoyun Hong, Alexander Fanaroff, Sophia Zhong, Kelvin N.V. Bush, Christopher Granger, Nicole Huber, Cynthia A. Jackevicius, Ingrid Kindipan, Shen Li, Paula Lewis, Asishana Osho, Jeffrey Sather, Stephen L. Sigal, Kathie Thomas, Zhao Ni, Jennifer L. Hall, Alice K. Jacobs, Abhinav Goyal

**Affiliations:** American Heart Association, Dallas, TX; Division of Cardiology, Perelman School of Medicine, University of Pennsylvania, Philadelphia, PA; Division of Cardiology, San Antonio Military Medical Center, San Antonio, TX; Duke University School of Medicine, Durham, NC; Emergency Medicine, Cumberland Healthcare Emergency Department, WI; Western University of Health Sciences, CA; VA Greater Los Angeles Healthcare System, University of Toronto, Canada ICES, Canada; Methodist Charlton Medical Center, TX; Five Rivers Medical Center, AR; Massachusetts General Hospital, MA; Trinity Health, ND; Heart and Vascular Institute at Titus, TX; Yale University School of Nursing, Orange, CT; Boston University Chobanian & Avedisian School of Medicine, MA; Division of Cardiology, Department of Medicine, Emory University School of Medicine, GA

## Abstract

**Background:** Cardiovascular disease (CVD) remains the leading cause of mortality worldwide, with acute myocardial infarction (AMI) contributing to over 100,000 deaths annually in the United States. Accurate risk stratification for in-hospital mortality is essential for guiding clinical decisions, improving outcomes, and optimizing hospital resources. However, existing models often rely on limited predictor sets, outdated data, and linear methods that may not reflect current clinical practice or complex interactions.

**Objective:** To develop and validate a contemporary in-hospital mortality risk model for AMI patients using modern statistical and machine learning approaches, incorporating clinical, demographic, and social determinants of health.

**Methods:** We utilized data from the American Heart Association (AHA) Get with The Guidelines®– Coronary Artery Disease (GWTG-CAD) Registry. Patients with AMI admitted between October 1, 2019, and December 31, 2022 (201,191 patients from 605 hospitals) were used to develop the in-hospital mortality prediction model. An independent validation cohort included 70,302 patients admitted in 2023. We incorporated 27 predictors, including demographics, comorbidities, vital signs, laboratory values, and social determinants of health. We assessed the models including a generalized linear mixed model (GLMM) and machine learning algorithms. We benchmarked against the legacy ACTION Registry–GWTG model. Subgroup and sensitivity analyses assessed model performance across sex, race/ethnicity, STEMI status, and time period.

**Results:** The LightGBM-based GWTG-CAD model achieved the highest discrimination (AUROC 0.874, 95% CI: 0.867–0.880) and demonstrated excellent calibration across all subgroups. Both GLMM and LightGBM outperformed the ACTION Registry–GWTG model, with LightGBM showing the most consistent performance in the 2023 validation cohort. Beyond traditional predictors, the inclusion of comorbidities, transportation methods, and community-level socioeconomic factors added predictive value.

**Conclusion:** The new GWTG-CAD risk model incorporates clinical, pre-hospital, and social factor predictors, along with machine learning methods, to enhance prediction of mortality in contemporary AMI patients.

## Introduction

Each year, acute myocardial infarction (AMI) affects approximately 1 million Americans and remains a leading cause of death in both the U.S. and worldwide.^1^ Accurate and well-calibrated models for predicting in-hospital mortality after AMI are critical for clinical decisions making, patient counseling, resource allocation, and hospital benchmarking. Notably, hospitals participating in the Get With The Guidelines® - Coronary Artery Disease (GWTG-CAD) Registry, one of the largest AMI registry in the U.S., rely on such mortality prediction models to support quality improvement and cross-hospital comparisons.^2,3^

While existing risk models were developed for in-hospital AMI mortality prediction, including the 2013 Acute Coronary Treatment and Intervention Outcomes Network (ACTION) Registry–GWTG model, ^4^ and GRACE Risk Score,^5^ these models were using older data and traditional statistical methods that may not reflect current clinical practice or capture nonlinear relationships among predictors. Shifts in population health, including the growing prevalence of obesity and diabetes ^6^ and changes in hospital systems further limit their relevance. Moreover, many models continue to use race as a proxy for risk, despite growing consensus that race is a social construct and that social determinants of health offer more precise and equitable predictors.^7,8^

To address these limitations, we developed a generalizable mortality risk model using both traditional statistical and machine learning models on contemporary data from the GWTG-CAD Registry. The goal of the study was to 1) test whether the inclusion of a broader set of clinical predictors and social determinants of health improves predictive performance; 2) determine whether the use of machine learning methods improves model performance compared with traditional linear models; and 3) develop a novel model, to be used for risk adjustment in quality improvement efforts.

## Methods

### Data source

The American Heart Association (AHA) GWTG-CAD Registry is a nationwide voluntary quality improvement program in the United States.^3^ Program information and data elements collected in the case report form are available at: https://www.heart.org/en/professional/quality-improvement. Participating hospitals upload clinical data of consecutive patients admitted with ST-elevation myocardial infarction (STEMI) or NSTEMI. Because data are used primarily at the local site for quality improvement, each participating hospital received either human research approval to enroll cases without individual patient consent under the Common Rule, or a waiver of authorization and exemption from subsequent review by their institutional review board (IRB). Advarra, the IRB for the American Heart Association, determined that this study is exempt from IRB oversight. The data collection and coordination for GWTG programs are managed by IQVIA (Parsippany, New Jersey).

### Study population

We included patients admitted to hospitals participating in the AHA GWTG-CAD registry between October 1, 2019, and December 31, 2022, as the primary cohort for model development and internal testing. To evaluate models’ robustness and validity, we identified a separate validation cohort comprising patients admitted between January 1, 2023, and December 31, 2023. Data preprocessing followed previously established methodology.^9^

Eligible patients presented to the hospital with signs, symptoms or complaints consistent with AMI (e.g., chest pain, tightness in chest or shortness of breath), and received a final diagnosis of AMI (STEMI or NSTEMI). AMI diagnosis was defined by either 1) Troponin I or Troponin T results > the upper limits of normal, or 2) ECG results consistent with AMI. Patients with missing data on sex, race/ethnicity, or discharge status, as well as those transferred out of participating hospitals, were excluded due to incomplete outcome data. The study population is shown in Figure S1.

### Outcome and Predictors

The outcome was in-hospital mortality, defined as death from any cause during the index hospitalization.

Predictors included variables recorded at the time of presentation, including demographics, medical history, comorbidities, electrocardiogram findings, initial vital signs, laboratory values, and socioeconomic status, with a total of 27 variables (**Table S1**), of which 9 were included in the legacy ACTION – GWTG risk score. Sex, race, and ethnicity were self-reported. Patient age at the time of diagnosis was calculated from the date of birth and the arrival date and time. Vital signs were values measured on first medical contact (FMC), by emergency medical services (EMS) for patients transported to the hospital by EMS, and in the emergency room for patients who transported themselves. Cardiac arrest prior to arrival indicates that the patient experienced an out-of-hospital cardiac arrest during pre-hospital care provided by EMS. Heart failure or cardiogenic shock on FMC was determined upon arrival at the hospital. Body mass index (BMI) was calculated using the first weight and height obtained upon hospital arrival. The initial serum creatinine and initial troponin value were defined as the first values acquired during the index hospitalization. STEMI on ECG was determined from provider documentation that the first or subsequent ECG findings were consistent with STEMI or a STEMI equivalent. Markers of socioeconomic status included insurance and social vulnerability index (SVI),^10^ which was determined based on patients residence. The SVI was calculated at the zip code level on the basis of 5-year estimates from the American Community Survey (2015– 2020) and linked to individual-level participant data in the GWTG cohorts. The SVI ranges from 0 to 1, with higher values indicating greater social vulnerability. To handle missing data, we applied median imputation for variables including initial serum creatinine, creatinine clearance, troponin levels, and SVI. We used multiple imputation with chained equations for BMI, systolic blood pressure (SBP), heart rate, smoking history, cardiogenic shock at FMC, cardiac arrest prior to arrival, and heart failure at FMC to preserve underlying distributions and relationships.

### Model development

We developed prediction models using two complementary methodologies. First, we applied a generalized linear mixed model, incorporating hospital site as a random effect to account for site-level variability in outcomes. Such a model offers the advantage of interpretability, allowing for the estimation of adjusted odds ratios and the inclusion of hospital-level effects. This modeling was also used in existing national benchmarks, such as the ACTION Registry–GWTG and GRACE models.^4^ Second, we implemented a range of machine learning models, including random forest, extreme Gradient Boosting Machine (XGBoost), and Light Gradient Boosting Machine (LightGBM). These tree-based ensemble methods are well-suited for clinical prediction tasks due to their ability to model complex, nonlinear interactions among variables. LightGBM and XGBoost, in particular, have demonstrated superior performance in a range of medical prediction studies.^11–13^

To benchmark our models, we also compared their performance to the ACTION Registry–GWTG model.^4^ For a fair comparison, we retrained the ACTION Registry–GWTG model using our training cohort, applying its published feature set and model structure. We then evaluated its performance on the validation dataset. Calibration performance was further examined across specific subgroups.

### Statistical Analysis

Summary statistics of baseline patient demographics and comorbidities, along with characteristics of the treating hospitals, were described using medians with interquartile range (IQR) and frequencies as appropriate. For descriptive analysis, we used the 2-proportion test for categorical values and used Kruskal-Wallis test for continuous variables. For generalized linear mixed models, we calculated odds ratios (ORs) with 95% CIs for each included variable and displayed results using a forest plot. For machine learning models, we used Shapley additive explanations (SHAP), to identify the most influential factors.

To evaluate model performance, the overall study cohort (2019–2022) was randomly split into training (80%) and validation (20%) sets using bootstrapping. Discrimination was assessed by the area under the receiver operating characteristic curve (AUROC), with 95% confidence intervals estimated via 100 bootstrap iterations. Calibration was evaluated both in the overall cohort and across predefined subgroups Several secondary analyses were conducted. We first evaluated the model performance across subgroups stratified by sex, race/ethnicity, STEMI vs. non-STEMI presentation, and year of diagnosis. Second, to assess generalizability in the post-pandemic era, we tested model performance using a separate cohort of patients admitted in 2023. Third, we examined the impact of including race as a predictor by developing models with and without race. These models were built using different sets of input variables—from basic demographics (e.g., age and sex) to more comprehensive inputs that included clinical history, laboratory results, and social factors such as insurance status and SVI. For each comparison, we evaluated differences in AUROC to determine whether the inclusion of race improved model performance.

All analyses were conducted using R (v4.2.0) and Python (v3.7.16) on the American Heart Association’s Precision Medicine Platform (https://pmp.heart.org). All *p* values are 2-sided. Because of the large size of the study population, statistical significance was defined as *p*<0.01.^14^

## Results

### Study population

The study cohort included 201,191 patients from 605 hospitals who were hospitalized with AMI (Table 1, Figure S1). Median age was 65 years (IQR: 57–75 years), and 32.7% of patients were female. 70.8% were Non-Hispanic White, 13.6% Non-Hispanic Black, and 8.1% Hispanic. In-hospital death occurred in 10,389 (5.2%) patients. Compared to patients who survived, patients who died were older (median age 72 vs. 65 years), more often female (39.1% vs. 32.4%), had diabetes mellitus (36.4% vs. 31.4%) and atrial fibrillation (12.9% vs. 7.4%). Patients who died were more likely to have had cardiogenic shock (31.1% vs. 2.8%) and cardiac arrest prior to hospital arrival (22.8% vs. 2.1%). The held-out validation cohort consisted of 70,302 patients hospitalized between January 1 and December 31, 2023, with similar distributions of demographic characteristics, comorbidities, and social factors compared to the overall study cohort (**Table S2)**.

**Table 1.** Baseline Characteristics.

| Variables | Patient Alive<br>(n = 190, 802) | Patient Died<br>(n = 10, 389) | Total<br>(n=201,191) | P value |
| --- | --- | --- | --- | --- |
| Age (years), Median (IQR) | 65 (18) | 72 (18) | 65 (18) | <0.001 |
| Female (n, %) | 61762<br>(32.37) | 4057 (39.05) | 65819<br>(32.71) | <0.001 |
| **Race/ethnicities (n, %)** |  |  |  |  |
| Non-Hispanic White | 135231<br>(70.88) | 7246 (69.75) | 142477<br>(70.82) | 0.014 |
| Non-Hispanic Black | 26099 (13.68) | 1269 (12.21) | 27368 | <0.001 |
|  |  |  | (13.60) |  |
| Hispanic | 15412 (8.08) | 910 (8.76) | 16322 (8.11) | 0.014 |
| BMI, Median (IQR) | 28.98 (7.28) | 27.90 (7.70) | 28.91 (7.32) | <0.001 |
| **Medical History (n, %)** |  |  |  |  |
| Diabetes Mellitus | 59916<br>(31.40) | 3780 (36.38) | 63696<br>(31.66) | <0.001 |
| Hypertension | 123435<br>(64.69) | 6811 (65.56) | 130246<br>(64.74) | 0.073 |
| Dyslipidemia | 98033<br>(51.38) | 4912 (47.28) | 102945<br>(51.17) | <0.001 |
| Smoking History | 64386<br>(33.74) | 2702 (26.01) | 67088<br>(33.35) | <0.001 |
| Currently on Dialysis | 4435 (2.32) | 544 (5.24) | 4979 (2.47) | <0.001 |
| Prior Myocardial<br>Infarction | 31187<br>(16.35) | 1541 (14.83) | 32728<br>(16.27) | <0.001 |
| Prior Percutaneous<br>Coronary Intervention | 37076<br>(19.43) | 1764 (16.98) | 38840<br>(19.31) | <0.001 |
| Prior CABG | 15005 (7.86) | 926 (8.91) | 15931 (7.92) | <0.001 |
| Atrial Fibrillation | 14110 (7.40) | 1343 (12.93) | 15453 (7.68) | <0.001 |
| Cerebrovascular<br>Diseases | 17007 (8.91) | 1369 (13.18) | 18376 (9.13) | <0.001 |
| Peripheral Arterial<br>Disease | 10012 (5.25) | 844 (8.12) | 10856 (5.4) | <0.001 |
| **Lab Values at Admission,**<br>Median (IQR) |  |  |  |  |
| Initial Serum Creatinine | 1.00 (0.40) | 1.40 (0.90) | 1.00 (0.4) | <0.001 |
| Creatinine Clearance | 80.39 (48.69) | 54.17 (46.96) | 80.39<br>(49.00) | <0.001 |
| Troponin | 0.19 (1.20) | 0.69 (5.48) | 0.2 (1.31) | <0.001 |
| STEMI on ECG (n, %) | 87355<br>(45.78) | 6783 (65.29) | 94138<br>(46.79) | <0.001 |
| Heart Rate at Admission,<br>Median (IQR) | 83 (27) | 87.67 (30) | 83 (27) | <0.001 |
| Systolic Blood Pressure at<br>Admission, mm Hg, Median<br>(IQR) | 150 (41) | 130 (45) | 149 (42) | <0.001 |
| Heart Failure FMC, (n, %) | 21803<br>(11.43) | 2776 (26.72) | 24579<br>(12.22) | <0.001 |
| Cardiogenic shock FMC (n,<br>%) | 5360 (2.81) | 3233 (31.12) | 8593 (4.27) | <0.001 |
| Cardiac arrest prior to<br>arrival (n, %) | 3942 (2.07) | 2367 (22.78) | 6309 (3.14) | <0.001 |
| Documented LVEF, % |  |  |  |  |
| <30% | 15770 (8.27<br>%) | 3138 (30.21<br>%) | 18908 (9.40<br>%) | <0.001 |
| 30 - 39% | 24086 (12.62) | 1733 (16.68) | 25819 (12.83) | <0.001 |
|  | %) | %) | %) |  |
| 40 - 49% | 37901 (19.86<br>%) | 1426 (13.73<br>%) | 39327 (19.55<br>%) | <0.001 |
| 50 - 59% | 62611 (32.81<br>%) | 1238 (11.92<br>%) | 63849 (31.74<br>%) | <0.001 |
| 60 - 69% | 40085 (21.01<br>%) | 811 (7.81 %) | 40896 (20.33<br>%) | <0.001 |
| >= 70% | 1830 (0.96 %) | 89 (0.86 %) | 1919 (0.95<br>%) | 0.320 |
| **Insurance (n, %)** |  |  |  |  |
| Medicaid | 23348<br>(12.24) | 1286 (12.38) | 24634<br>(12.24) | 0.6791 |
| Medicare | 54762<br>(28.70) | 4119 (39.65) | 58881<br>(29.27) | <0.001 |
| Private/VA/CHAMPUS | 91622<br>(48.02) | 3881 (37.36) | 95503<br>(47.47) | <0.001 |
| Self-pay/No Insurance | 12433 (6.52) | 602 (5.79) | 13035 (6.48) | 0.004 |
| Not Documented/UTD | 8637 (4.53) | 501 (4.82) | 9138 (4.54) | 0.166 |
| **Transportation (n, %)** |  |  |  |  |
| EMS – Air | 1348 (0.71) | 111 (1.07) | 1459 (0.73) | <0.001 |
| EMS – Ground<br>Ambulance | 75423<br>(39.53) | 6350 (61.12) | 81773<br>(40.64) | <0.001 |
| Walk-in | 70591 | 1777 (17.10) | 72368 | <0.001 |
|  | (37.00) |  | (35.97) |  |
| Transferred from another facility | 43440<br>(22.77) | 2151 (20.70) | 45591<br>(22.66) | <0.001 |
| SVI (0-1), Median (IQR) | 0.43 (0.40) | 0.43 (0.41) | 0.43 (0.40) | <0.001 |

Values for continuous variables are median (interquartile range).
Abbreviations: IQR, interquartile range; BMI, body mass index; MI, myocardial infarction; PCI, percutaneous coronary intervention; CABG, coronary artery bypass grafting; ECG, electrocardiogram; STEMI, ST-segment elevation myocardial infarction; FMC, first medical contact; EMS, emergency medical services; LVEF, Left Ventricular Ejection Fraction; SVI, social vulnerability index; VA, Veterans Affairs; CHAMPUS, Civilian Health and Medical Program of the Uniformed Services; UTD, unable to determine.

**Table 2.** Model performance on total cohort (AUROC).

| **Models** | **ACTION Registry – GWTG** | **GLMM** | **Random Forest** | **XGBoost** | **LightGBM (GWTG-CAD model)** |
| --- | --- | --- | --- | --- | --- |
| AUROC | 0.859 (0.852 - 0.867) | 0.865 (0.859 - 0.872) | 0.857 (0.849 - 0.865) | 0.865 (0.858 - 0.871) | 0.874 (0.867 - 0.880) |

### Associations Between Baseline Characteristics and in-Hospital Mortality

Figure 1 presents the associations between baseline clinical variables and in-hospital mortality, as estimated by a generalized linear mixed model. The variables with the highest odds ratios were cardiac arrest prior to arrival (OR 6.29, 95% CI: 5.78–6.84) and cardiogenic shock at first medical contact (OR 4.65, 95% CI: 4.32–5.01), indicating the strongest associations with in-hospital mortality. Other clinical predictors significantly associated with increased mortality included age, STEMI on ECG, and heart failure at presentation. Several clinical variables not included in previous models were also significantly associated with in-hospital mortality, including diabetes mellitus, atrial fibrillation, cerebrovascular disease, and prior coronary artery bypass grafting. Although the median and IQR of SVI were nearly identical between survivors and non-survivors (0.43 [0.40] vs. 0.43 [0.41]), the model revealed that even within this narrow range, incremental increases in social vulnerability were significantly associated with higher odds of mortality (OR = 1.18, CI 1.06-1.32). Transportation to the hospital by ground EMS (OR = 1.49, 95% CI 1.40–1.60) or transfer from another facility (OR = 1.38, 95% CI 1.28–1.50) were associated with higher in-hospital mortality compared with patients who transported themselves to the emergency room.

**Figure 1.**
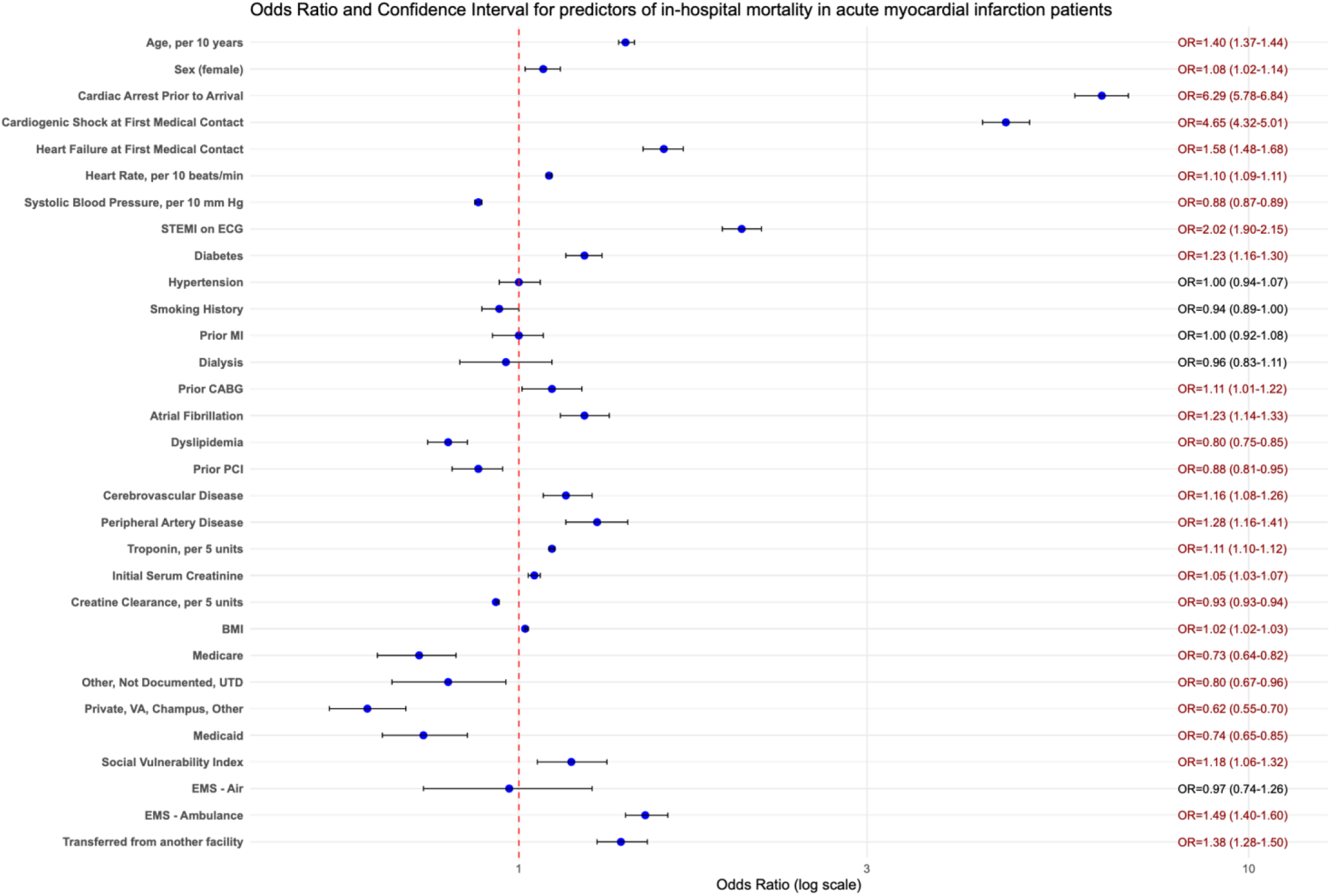
Odds Ratios (OR) and 95% Confidence Intervals (CI) for Predictors of In-Hospital Mortality in Acute Myocardial Infarction Patients (GLMM model)

Including race as a predictor provided minimal incremental improvement in model performance across varying levels of model complexity (Table S3). In a model with only demographic and clinical severity variables, the discrimination was modest (AUROC = 0.8186). Model performance improved with the addition of medical history, laboratory values, vital signs, and social determinants of health, achieving an AUROC of 0.8685. When race was included in the fully adjusted model, the increase in AUROC was less than 0.001, indicating that race contributed little additional predictive value when comprehensive clinical and social factors were already accounted for.

### Model Performance Characteristics

Among all modes that we tested, the highest median AUROC was achieved by the LightGBM model, with a median AUROC of 0.874 (95% CI, 0.867–0.880) (Table 1). The generalized linear mixed model achieved a median AUROC of 0.865 (95% CI, 0.859–0.872), similar to the performance of models built using random forest and XGBoost models. In the separate validation cohort of patients (n=59,730 patients), the LightGBM’s performance remained stable (AUROC 0.872), outperforming the legacy ACTION Registry – GWTG model (AUROC 0.855).

Calibration plots (Figure 2) showed that LightGBM and Random Forest provided the best-calibrated predictions, with predicted mortality closely matching observed mortality across deciles. In the separate validation cohort, the legacy ACTION Registry – GWTG model and the generalized linear mixed model overestimated mortality across high-risk patients.

**Figure 2.**
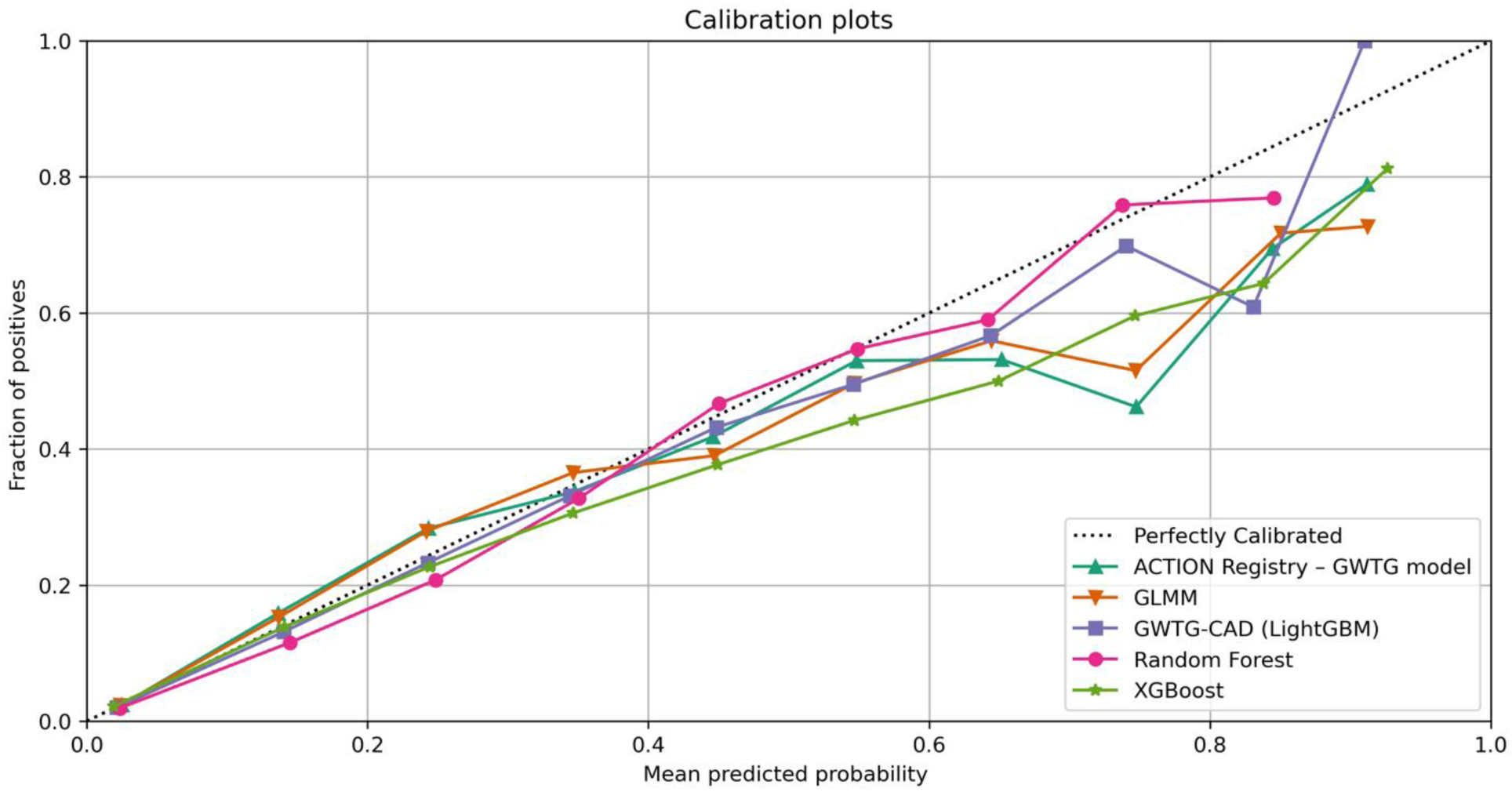
Calibration of prediction models using the held-out test set.

### Sensitivity analysis

In subgroup analyses (Table 3) across all sex, race/ethnicity, clinical presentation, and calendar year subgroups, LightGBM achieved the highest AUROC, indicating superior discriminative performance. In calibration subgroup analyses (Figure 3), both LightGBM maintained consistent calibration, whereas the ACTION Registry – GWTG model and the generalized linear mixed model each overestimated risk in the highest risk patients, such as female, and racial/ethnic minority groups. Among patients with NSTEMI, the LightGBM model shows the best calibration, with slightly underprediction for high risk. The ACTION Registry – GWTG model and generalized linear mixed performs well at low-to-mid risk but overestimates risk between 0.7–0.8. These patterns may reflect the greater clinical heterogeneity and complexity typically observed in NSTEMI compared to STEMI patients.^15^

**Figure 3.**
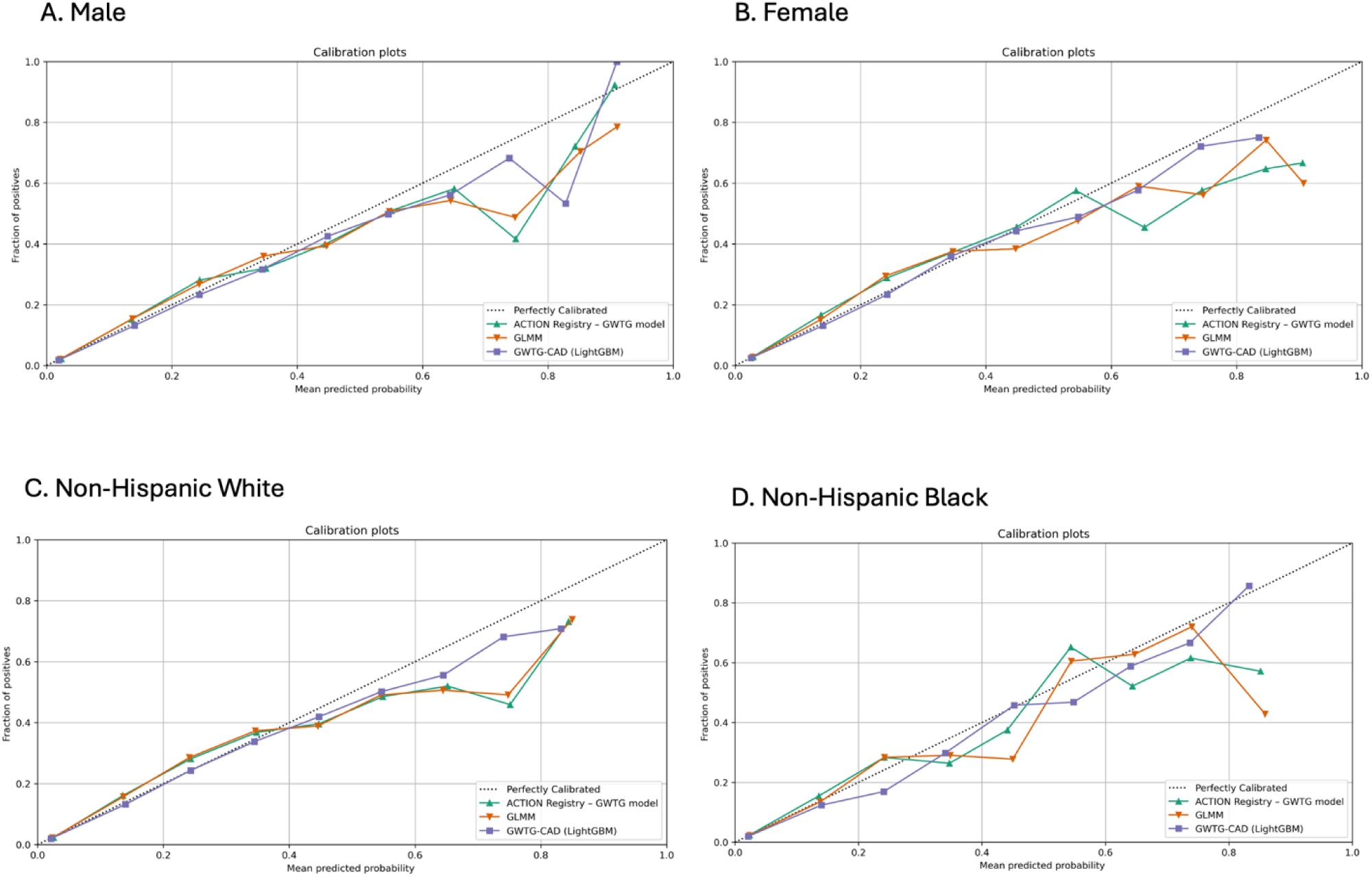
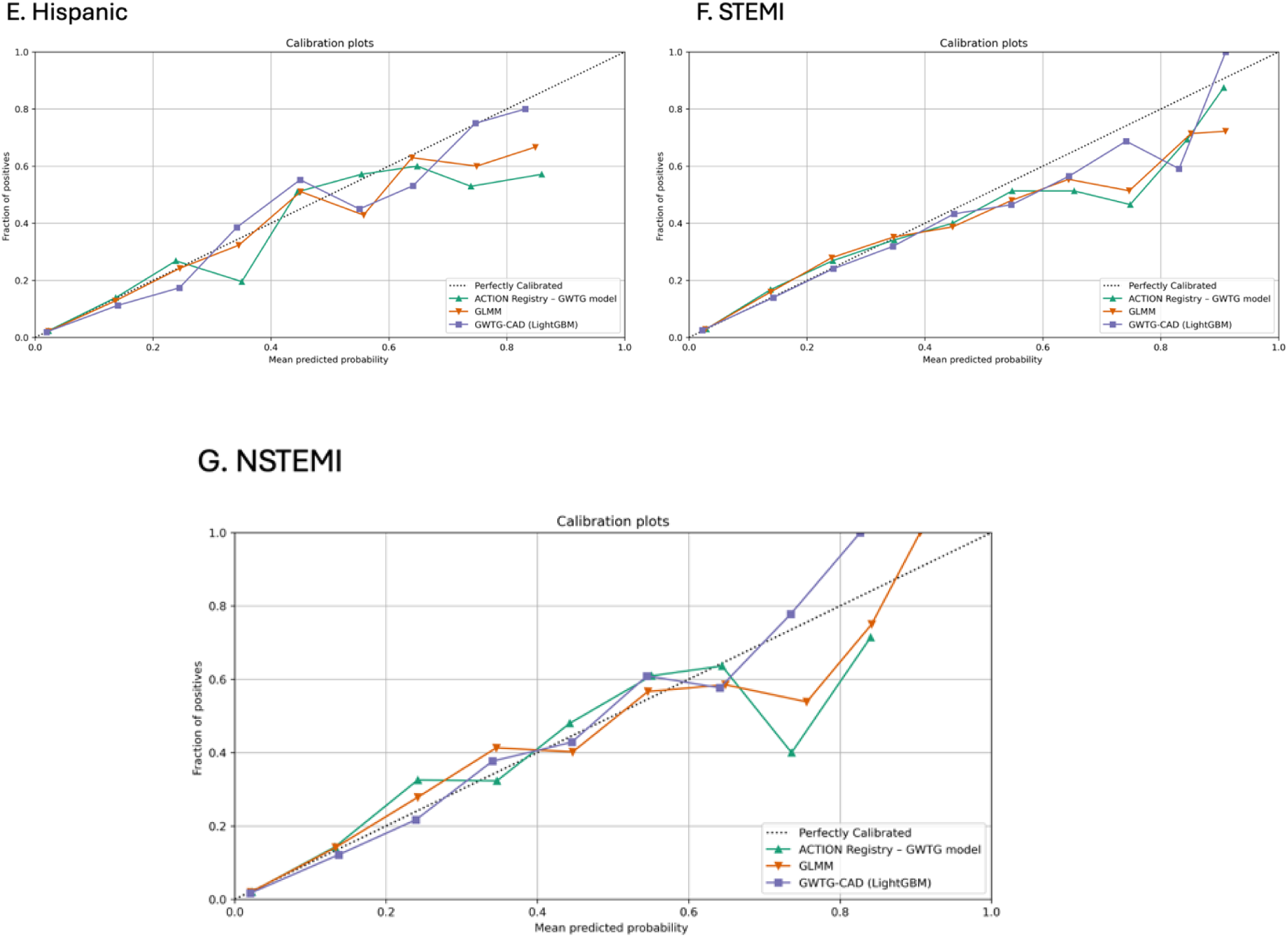
Calibration of prediction models on subgroup.

**Table 3.**
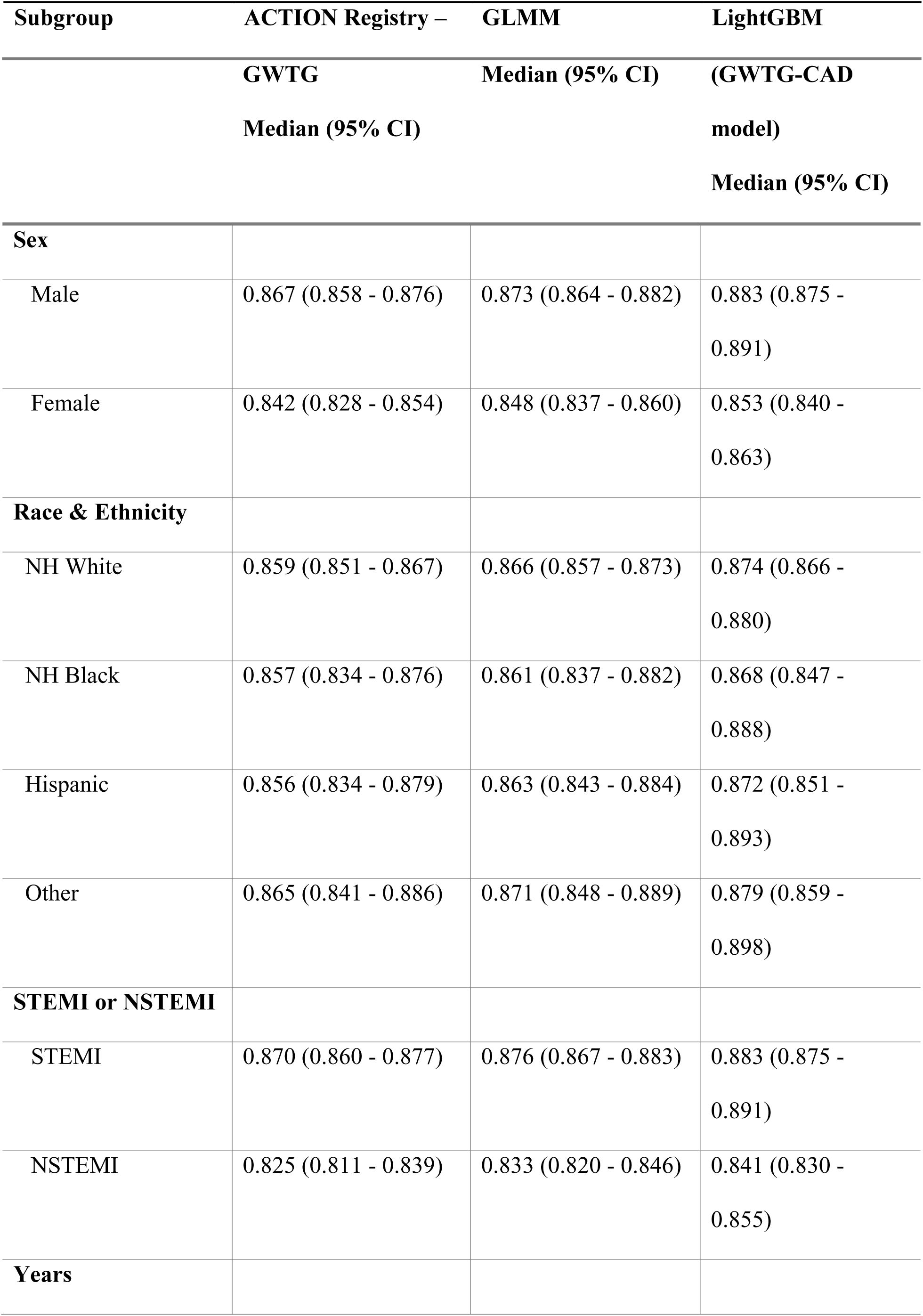
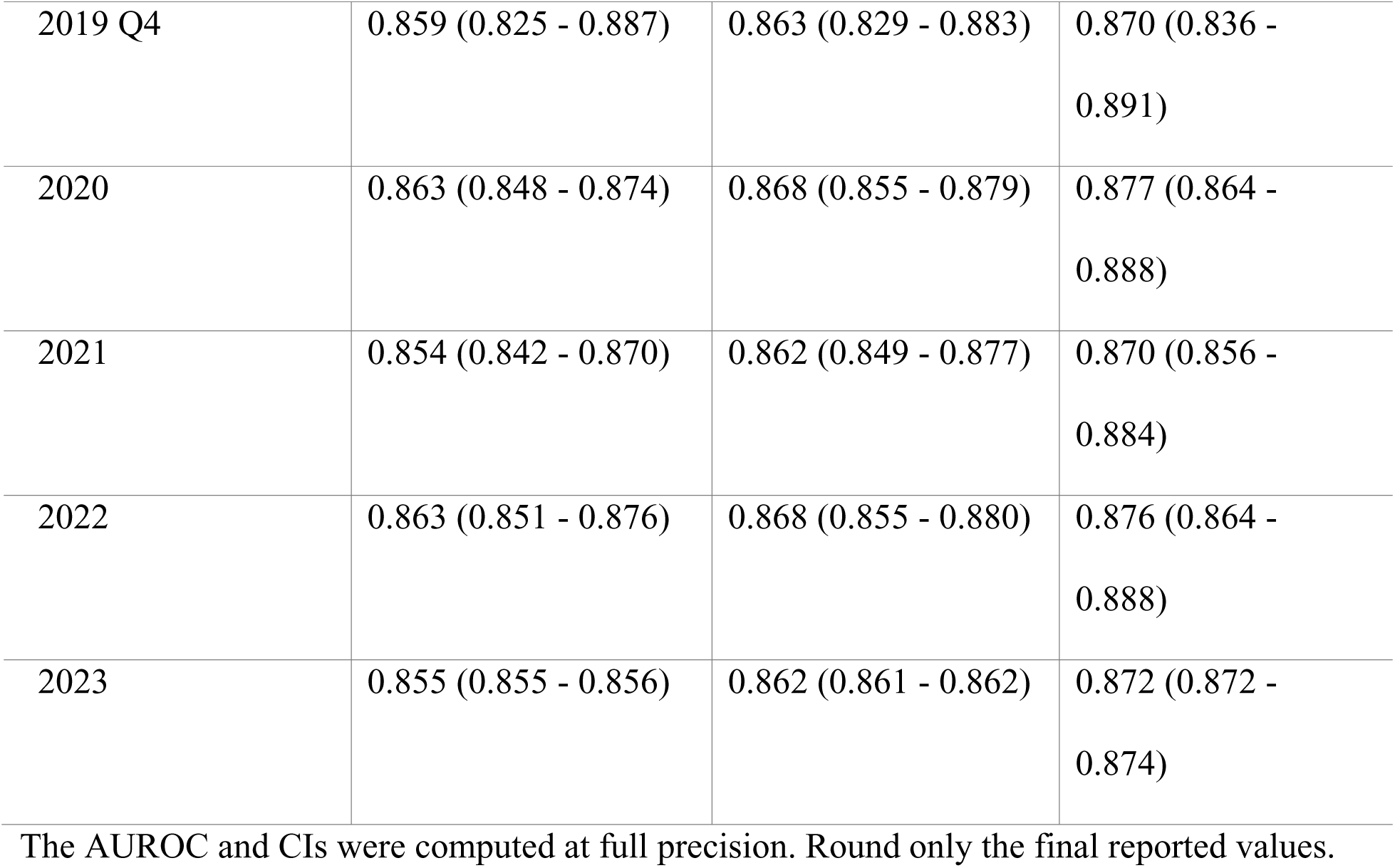
Discrimination (AUROC) on subgroup stratified by sex, race & ethnicity, STEMI or NSTEMI, years group.

| **Subgroup** | **ACTION Registry –** | **GLMM** | **LightGBM** |
| --- | --- | --- | --- |

|  | **GWTG** | **Median (95% CI)** | **(GWTG-CAD** |
| --- | --- | --- | --- |
|  | **Median (95% CI)** |  | **model)** |
|  |  |  | **Median (95% CI)** |
| **Sex** |  |  |  |
| Male | 0.867 (0.858 - 0.876) | 0.873 (0.864 - 0.882) | 0.883 (0.875 - 0.891) |
| Female | 0.842 (0.828 - 0.854) | 0.848 (0.837 - 0.860) | 0.853 (0.840 - 0.863) |
| **Race & Ethnicity** |  |  |  |
| NH White | 0.859 (0.851 - 0.867) | 0.866 (0.857 - 0.873) | 0.874 (0.866 - 0.880) |
| NH Black | 0.857 (0.834 - 0.876) | 0.861 (0.837 - 0.882) | 0.868 (0.847 - 0.888) |
| Hispanic | 0.856 (0.834 - 0.879) | 0.863 (0.843 - 0.884) | 0.872 (0.851 - 0.893) |
| Other | 0.865 (0.841 - 0.886) | 0.871 (0.848 - 0.889) | 0.879 (0.859 - 0.898) |
| **STEMI or NSTEMI** |  |  |  |
| STEMI | 0.870 (0.860 - 0.877) | 0.876 (0.867 - 0.883) | 0.883 (0.875 - 0.891) |
| NSTEMI | 0.825 (0.811 - 0.839) | 0.833 (0.820 - 0.846) | 0.841 (0.830 - 0.855) |
| **Years** |  |  |  |
| 2019 Q4 | 0.859 (0.825 - 0.887) | 0.863 (0.829 - 0.883) | 0.870 (0.836 -<br>0.891) |
| 2020 | 0.863 (0.848 - 0.874) | 0.868 (0.855 - 0.879) | 0.877 (0.864 -<br>0.888) |
| 2021 | 0.854 (0.842 - 0.870) | 0.862 (0.849 - 0.877) | 0.870 (0.856 -<br>0.884) |
| 2022 | 0.863 (0.851 - 0.876) | 0.868 (0.855 - 0.880) | 0.876 (0.864 -<br>0.888) |
| 2023 | 0.855 (0.855 - 0.856) | 0.862 (0.861 - 0.862) | 0.872 (0.872 -<br>0.874) |

The AUROC and CIs were computed at full precision. Round only the final reported values.

For clarity in figures and tables, we refer to the LightGBM-based model as the GWTG-CAD risk model, given its superior performance in both discrimination and calibration.

### Feature importance of machine learning model

To enhance model interpretability, we used SHapley Additive exPlanations (SHAP) to visualize how the final GWTG-CAD model generated mortality predictions (Figure 4). Overall, age was the strongest predictor, followed by systolic blood pressure, heart rate, and STEMI presentation. The beeswarm plot revealed that lower systolic blood pressure values were strongly associated with increased mortality risk, while mid-to-high values were associated with decreased risk.

**Figure 4.**
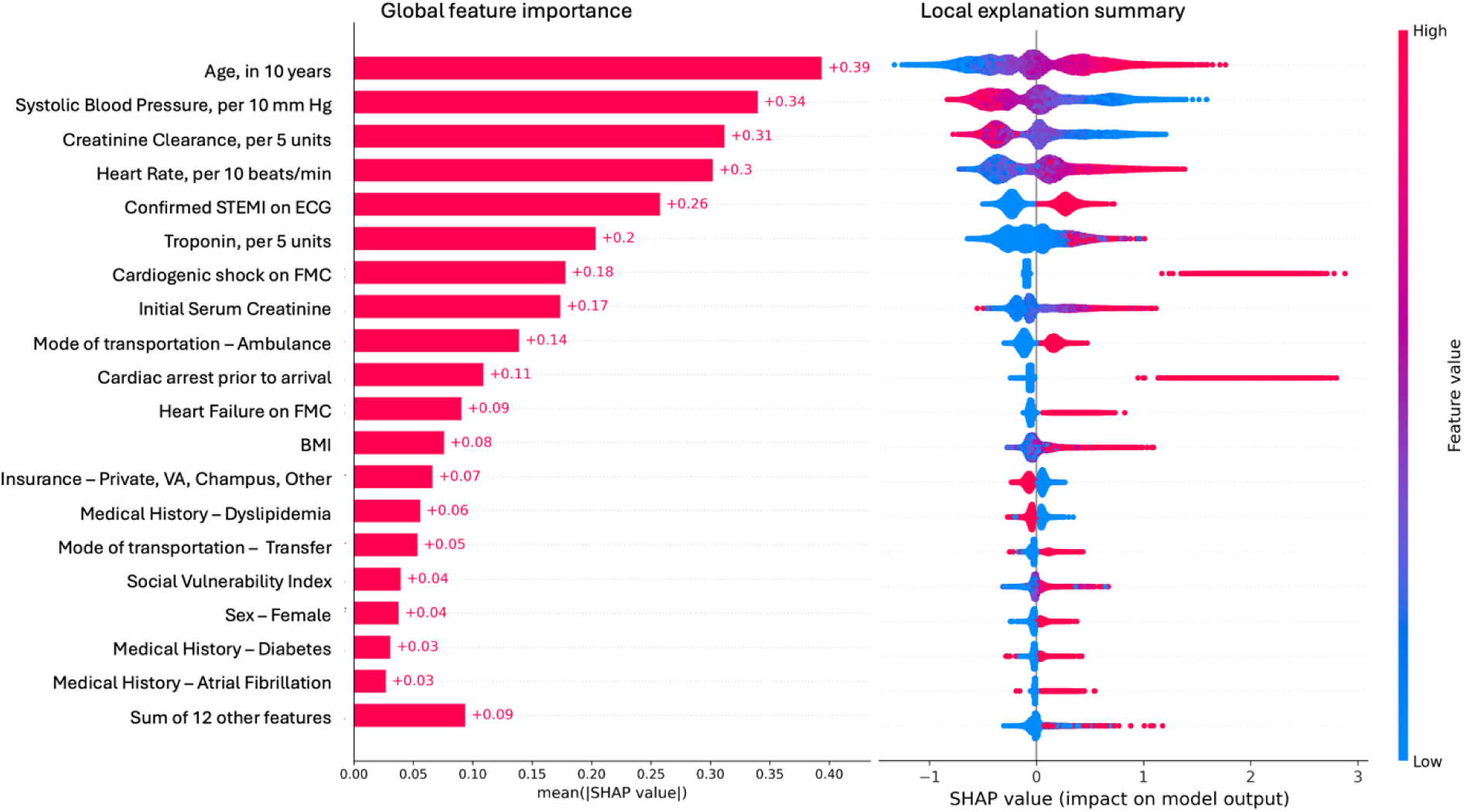
Global Feature Importance and Local Explanation Summary for Predicting In-Hospital Mortality in AMI Patients Using SHAP Values in the GWTG-CAD model. The left side is the global importance ranking of features that represents the average influence of the feature across the entire dataset. The right side of beeswarm plots show how a feature impacts individual predictions.

Renal function markers including initial serum creatinine and creatinine clearance were also prominent contributors, underscoring the importance of renal dysfunction in acute mortality risk. Features such as cardiac arrest, cardiogenic shock, and heart failure at presentation, had high SHAP values for certain patients but lower predictive importance due to their low prevalence in the overall cohort. Social and system-level factors, including SVI, insurance type, and mode of arrival (e.g., EMS transport), were also retained as influential, highlighting the value of incorporating social context into risk prediction models.

## Discussion

Accurate in-hospital risk prediction for patients with AMI remains a critical endeavor with significant implications for clinical decision-making and hospital quality assessment. The dynamic nature of clinical practice, evolving patient demographics, and shifting social determinants of health necessitate the continuous development and validation of contemporary risk stratification tools. This study addresses this need by using contemporary nationwide data, state-of-the-art machine learning methods, and a broad set of prospectively-collected prediction variables to develop the novel risk model.

Our study offers several key advancements over existing benchmarks. First, we incorporated an expanded set of clinical variables alongside community-level socioeconomic factors, allowing for a more comprehensive assessment of risk. Second, we rigorously evaluated both traditional statistical methods (generalized linear mixed model) and advanced machine learning approaches, with GWTG-CAD model demonstrating superior performance in both discrimination and calibration. Third, we conducted a robust evaluation of model performance across various demographic and clinical subgroups, as well as over time, including during the COVID-19 pandemic.

The GWTG-CAD model consistently exhibited good to excellent discrimination, achieving an AUROC of 0.874 in the validation cohort, outperforming the generalized linear mixed model and the legacy model, which achieved an AUROC of 0.859 in our test set. This superior performance was maintained across demographic and clinical subgroups, highlighting the potential for more equitable risk assessment. Furthermore, the GWTG-CAD model demonstrated better calibration than the generalized linear mixed model, including across subgroups, and its calibration remained stable over time, underscoring its generalizability to current clinical settings.

Enhanced predictive performance of the novel GWTG-CAD model compared with previous models can be explained both by its use of a greater set of predictive variables and use of modern statistical methods. A generalized linear mixed model using a larger set of variables had enhanced discrimination (though not calibration) compared with the legacy model, and the novel machine learning model had still greater discrimination. This emphasizes the need for regular updates to risk prediction models to maintain accuracy and equity. While the ACTION Registry–GWTG model remains valuable as a historical benchmark, our study highlights the potential of our newly developed models to provide more accurate risk estimates in the current era.

Our analysis also provides insights into the predictive value of various factors. Beyond well-established predictors such as age, hemodynamic instability, and clinical severity at admission, we found that comorbidities, transportation method, and community-level socioeconomic factors contributed added value to risk prediction. Interestingly, the inclusion of self-reported race did not enhance model performance once all clinical and socioeconomic predictors were accounted for. This suggests that race-associated differences in outcomes may be mediated through the measurable clinical and socioeconomic factors already captured in our models.

We acknowledge several limitations in our study. Only patients presenting to facilities participating in GWTG-CAD were included. Participation in this registry is voluntary and thus may include higher volume hospitals providing higher-quality care than hospitals not included. This may limit the generalizability of our findings to hospitals that did not participate in GWTG-CAD. Additionally, the performance of our models varied slightly across subgroups, with minor under-prediction or over-prediction observed for Non-Hispanic Black and Hispanic patients, warranting further investigation into potential sources of these discrepancies. Finally, we relied on county-level SVI to estimate socioeconomic status, which may not fully capture individual-level variations.

Despite these limitations, this study offers an updated and robust risk prediction model for in-hospital mortality following AMI. Future research should focus on validation of these models in external cohorts and their integration into electronic health record (EHR) systems to facilitate real-time risk assessment. Furthermore, assessing the impact of these models on real-world clinical decisions and outcomes is crucial to translate these findings into tangible improvements in patient care. The accurate calibration of our models is particularly important in the context of risk-adjusted hospital performance metrics, which are increasingly used for public reporting and may have financial implications.

In conclusion, this study presents a novel and validated in-hospital risk prediction model – the GWTG-CAD model—for AMI that leverages contemporary data, expanded clinical and socioeconomic variables, and advanced machine learning techniques. The superior performance and calibration of the GWTG-CAD model, along with the insights gained into the contribution of various risk factors, represent a significant advancement in the field and hold promise for enhancing the accuracy and equity of risk stratification in AMI patients.

## Data Availability

The Precision Medicine Platform is a cloud-based system that allows researchers to collaborate and analyze large datasets from any computer in the world using a secure environment and the power of machine learning. Within the research interface, users have access to assorted datasets, including the industry-changing Get With The Guidelines® registry data to accelerate findings into impactful discoveries. Researchers must have an approved research proposal to access Get With The Guidelines® registry data.

https://www.heart.org/en/professional/quality-improvement/get-with-the-guidelines/get-with-the-guidelines-coronary-artery-disease

